# Longitudinal MAP-MRI-based Assessment of Tissue Microstructural Alterations in Acute mTBI

**DOI:** 10.64898/2026.04.06.26350074

**Authors:** Mihika Gangolli, Neil J. Perkins, Luca Marinelli, Peter J. Basser, Alexandru V. Avram

## Abstract

**BACKGROUND:** Mild traumatic brain injury (mTBI) is a signature injury in civilian and military populations that remains invisible to detection by conventional radiological methods. Diffusion MRI has been identified as a potential clinical tool for revealing subtle microstructural alterations associated with mTBI.

**OBJECTIVE:** This study evaluates whether a comprehensive and powerful diffusion MRI (dMRI) technique called mean apparent propagator (MAP) MRI can detect sequelae of mTBI.

**METHODS:** We analyzed data from 417 participants of the GE/NFL prospective mTBI study which included 143 matched controls (mean age, 21.9 ± 8.3 years; 76 women) and 274 patients with acute mTBI and GCS ≥13 (mean age, 21.9 ± 8.5 years; 131 women). All participants underwent MRI exams at up to four visits including structural high-resolution T_1_W, T_2_W, FLAIR-T_2_W, and dMRI, in addition to clinical assessments of post-concussive physical symptoms (RPQ-3), psychosocial functioning and lifestyle symptoms (RPQ-13), and postural stability (BESS). The dMRI data for each subject were co-registered across all visits and analyzed using the MAP-MRI framework to measure and map the distribution of net microscopic displacements of diffusing water molecules in tissue and ultimately compute the microstructural MAP-MRI tissue parameters including propagator anisotropy (PA), Non-Gaussianity (NG), return-to-origin probability (RTOP), return-to-axis probability (RTAP), and return-to-plane probability (RTPP). We quantified voxel-wise and region-of-interest (ROI)-based changes in these parameters across all four visits.

**RESULTS:** MAP-MRI parameter values were within the expected ranges and showed relatively little variation across visits. We found no significant differences in the longitudinal trajectories of these parameters between mTBI patients and controls. At acute post-injury timepoints, RPQ-3 and RPQ-13 scores were increased in mTBI patients relative to controls, while BESS scores were not significantly different between groups. Analysis of dMRI metrics and clinical mTBI markers showed significant correspondence between MAP-MRI metrics in cortical gray matter, caudate and pallidum and BESS scores.

**CONCLUSION:** We developed and tested a state-of-the-art quantitative image processing pipeline for sensitive analysis and detection of subtle tissue changes in longitudinal clinical diffusion MRI data. The absence of a significant statistical difference between populations in the dMRI parameters in this study suggests that the mTBI corresponded to acute post-injury clinical symptoms but that the injury was not severe enough to cause detectable microstructural damage/alterations, and that increased diffusion sensitization combined with improved analysis techniques may be needed.

**CLINICAL IMPACT:** These findings suggest that acute mTBI (GCS≥13) may not be detectable with diffusion MRI.

**TRIAL REGISTRATION:** ClinicalTrials.gov NCT02556177

## Introduction

Mild traumatic brain injury (mTBI) is prevalent in both civilian and military populations, representing a substantial public health burden. Although classified as “mild,” mTBI is characterized by subtle microstructural alterations, including cellular edema, gliosis, and traumatic axonal injury (Sturdivant et al. 2016; Ogino et al. 2022). Up to 50% of patients report persistent neurological symptoms and reduced quality of life in the months to years after injury (Mcgeown et al. 2025). Despite these clinical sequelae, conventional imaging findings with CT or structural MRI in mTBI patients are typically radiologically normal as these techniques are primarily sensitive to macroscopic abnormalities such as hemorrhage or contusions (Mayer et al. 2020). Consequently, many mTBI cases go undetected radiologically, and imaging findings often fail to correspond to symptom severity. This discordance likely reflects the spatial heterogeneity of tissue injury, inter-individual variability in pathophysiological responses, and the evolving temporal dynamics of post-traumatic processes. These limitations underscore the need to develop sensitive and specific imaging biomarkers capable of detecting subtle microstructural injury to inform mTBI patient care.

Diffusion MRI (dMRI) is a promising candidate for mTBI diagnosis because it efficiently and non-invasively probes tissue microstructure throughout the entire brain. However, diffusion findings in mTBI have been inconsistent. Reported abnormalities vary in direction, magnitude, and spatial distribution, which may reflect heterogeneity in injury location, differences in inflammatory and reparative responses, and variability in acquisition protocols and data processing approaches. Many prior studies have relied on tensor-based metrics or atlas-based group comparisons, which require spatial normalization across subjects and large cohort sizes. Such approaches are inherently limited in mTBI, where lesions are subtle, spatially heterogeneous, and patient-specific. Furthermore, cross-sectional designs with only one or two post-injury time points may obscure dynamic microstructural changes that evolve over days to weeks.

Longitudinal MRI studies could provide valuable insights into the progression of subtle spatiotemporal changes in radiological outcomes that are typically obscured due to cross-sectional study designs (Echlin, Rahimi, and Wojtowicz 2021). Previous quantitative longitudinal diffusion imaging studies of mTBI have used approaches to register subject data to a common atlas or template in order to identify regions with altered tissue diffusivity properties (Palacios et al. 2022). This approach requires large cohort sizes and typically consists of data acquired at two time points after injury, but findings are limited by the interpolation of the relatively lower spatial resolution of the diffusion data in a heterogeneous subject population. An alternative approach with data acquired at more time points would provide increased statistical power when performing within-subject registration while preserving the ability to identify patient-specific changes in tissue diffusion after mTBI. Longitudinal imaging studies offer a complementary strategy that may enhance sensitivity to subtle injury-related changes. Experimental and clinical studies have demonstrated that molecular and cellular markers of injury—including inflammatory mediators and indicators of tissue remodeling—exhibit distinct temporal trajectories in the days and weeks following mTBI (Adrian, Marten, Salla, et al. 2016). Capturing analogous temporal patterns in tissue microstructure may provide diagnostically meaningful information that is not apparent in cross-sectional analyses. Within-subject longitudinal designs reduce inter-individual variability and may enable detection of dynamic microstructural alterations that would otherwise be obscured or washed-out in group-level comparisons.

Mean apparent propagator (MAP) MRI (Ö zarslan et al. 2013) provides a comprehensive clinically feasible framework (Avram et al. 2016) for characterizing diffusion by estimating the full three-dimensional net displacement probability density function of water molecules. Unlike model-specific approaches, MAP-MRI removes the dependence on imaging and acquisition parameters and directly reconstructs the diffusion propagator, yielding a family of complementary quantitative parameters that includes the return-to-origin probability (RTOP), return-to-axis probability (RTAP), return-to-plane probability (RTPP), propagator anisotropy (PA), Non-Gaussianity (NG), and mean squared displacement (MSD). These MAP-MRI parameters complement the DTI metrics (Avram, Hutchinson, and Basser 2017), reflecting important features of tissue microstructure and cytoarchitecture accessible at stronger diffusion sensitizations (Avram, Saleem, Komlosh, et al. 2022; Saleem, Avram, Yen, et al. 2023; Pas et al. 2024). At long diffusion times, RTOP, RTAP, and RTPP have been linked to geometric descriptors of restricted environments, reflecting mean pore volume, cross-sectional area, and length within a voxel (Avram and Basser 2014; Zucchelli et al. 2016; Komlosh et al. 2017). These properties make MAP-MRI particularly well-suited for detecting subtle, spatially heterogeneous microstructural alterations (Avram, Bernstein, Irfanoglu, et al. 2019).

MAP-MRI metrics have demonstrated high reproducibility in clinical acquisitions (Avram et al. 2016) and have shown associations with clinical outcomes in neurological disorders, such as Alzheimer’s disease (Spotorno et al. 2022), Parkinson’s disease (Le et al. 2020), ischemic stroke (Boscolo Galazzo et al. 2018), cancer (Wang et al. 2022; She et al. 2023), epilepsy (Chen et al. 2019), traumatic brain injury (Hutchinson, Schwerin, Avram, et al. 2018), and chronic traumatic encephalopathy (Gangolli et al. 2023). A correlation of MAP-MRI data and pathological markers in tissue specimens with a post-mortem diagnosis of chronic traumatic encephalopathy showed correlations of phosphorylated tau with Non-Gaussianity (NG) and astrogliosis with RTOP (Gangolli et al. 2023). Previous investigation of clinically acquired MAP-MRI data in a pilot cohort of mTBI subjects has assessed the test-retest variability of neurologically healthy controls and demonstrated the feasibility of evaluating the spatiotemporal trends of longitudinal changes after mTBI (Gangolli et al. 2025).

In this study, we aim to characterize the spatiotemporal evolution of tissue microstructural alterations following acute mTBI using a within-subject longitudinal design. By quantifying changes in the diffusion propagator and associated MAP-MRI metrics over time, we seek to establish sensitive imaging biomarkers of injury-related microstructural alterations that may ultimately support diagnosis, treatment, and improve patient management.

## Methods

### Subject Selection

This retrospective longitudinal analysis was performed using a pilot subset of data previously acquired between 2015 and 2018 as part of the Advanced MRI Applications for Mild Traumatic Brain Injury – Phase 2 multicenter study (ClinicalTrials.gov identifier: NCT02556177) (Tanwar et al. 2025). The parent study was conducted at seven academic medical centers (Hospital for Special Surgery, New York, NY; Houston Methodist, Houston, TX; University of California, San Francisco, CA; University of Pittsburgh Medical Center, Pittsburgh, PA; Medical College of Wisconsin, Milwaukee, WI; University of Miami, Miami, FL; University of California, San Diego). Each participating site obtained Institutional Review Board approval, and written informed consent was obtained from all participants or their legal guardians.

Patients with mTBI were referred from concussion clinics at each site. Eligibility criteria included age 15–50 years, Glasgow Coma Scale (GCS) score ≥13 at presentation, and imaging within 10 days of injury. mTBI diagnosis was established according to the Zurich consensus statement (McCrory et al. 2013) and the American Congress of Rehabilitation Medicine (ACRM) criteria (Mild Traumatic Brain Injury Committee of the Head Injury Interdisciplinary Special Interest Group of the American Congress of Rehabilitation Medicine 1993).

Neurologically healthy controls were recruited at each site and were required to meet age criteria (15–50 years). Controls with a history of mTBI were required to be *>*6 months post-injury.

Exclusion criteria for all participants included: history of moderate-to-severe TBI (GCS ≤13), epilepsy with recurrent seizures within the prior 10 years, contraindication to MRI, structural brain abnormality on prior MRI, primary Axis I or II psychiatric disorder, or history of substance abuse.

### MR Data Acquisition

Supplementary Figure 1 shows the timeline of data acquisition and the number of participants per site. Patients with mTBI were scanned at four post-injury visits: (Visit_1_ – *<*72 hours; Visit_2_ – 5-10 days; Visit_3_ – 12-16 days; Visit_4_ – 83-97 days) post-trauma. MRI data were acquired on 3T GE Signa MR750 systems (GE Healthcare) using 32-channel RF head coils (Nova Medical). The acquisition parameters for each MRI dataset are shown in Table 2. During each post-injury visit, total balance error scoring (BESS) on firm and foam surfaces and Rivermead post-concussion questionnaire (RPQ-3 and RPQ-13) were used to assess postural stability and self-reported post-concussive symptoms, respectively.

### MR Image Processing

#### Data Quality Control and Exclusion

Figure 1 shows the data analysis pipeline. Automated outlier detection was performed to identify scans affected by structural abnormalities or field inhomogeneities. The algorithm quantified voxel count and intensity distributions across axial, coronal, and sagittal planes and compared voxel-wise histograms to modeled control bounds. Scans were excluded if 10 percent of voxels were affected by artifacts or structural abnormalities.

**Figure 1:**
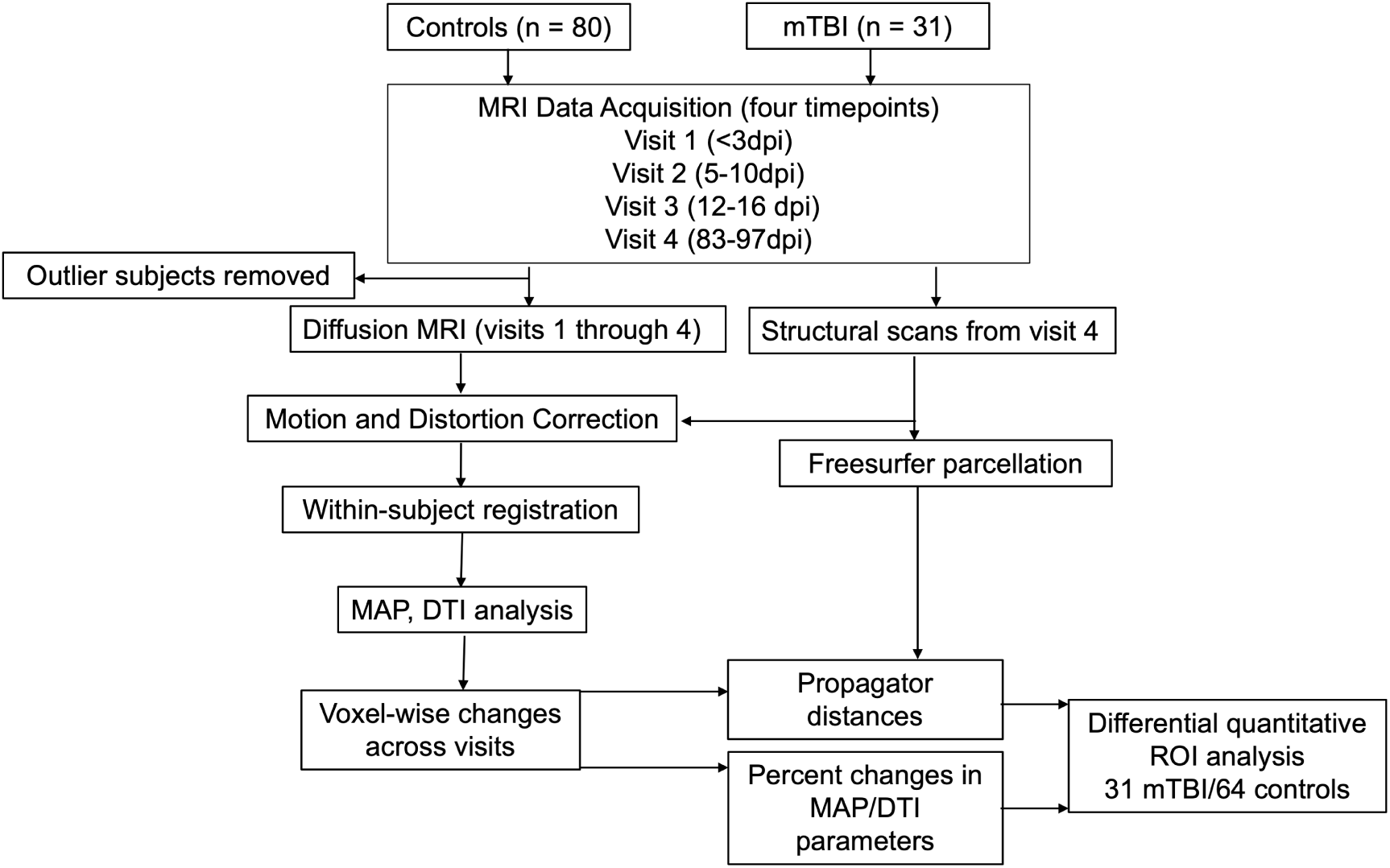
Longitudinal MAP-MRI acquisition and analysis pipeline for evaluation of spatiotemporal changes following mTBI. MRI data from control and mTBI subjects were acquired across six sites at four post-injury timepoints. The diffusion MRI datasets were registered within-subject to data from the final visit. ROIs were used to summarize spatial patterns of voxel-based differences in the diffusion propagator and diffusion MRI metrics. Abbreviations: mTBI – mild traumatic brain injury; MAP – mean apparent propagator; DTI – diffusion tensor imaging; ROI – region of interest.

#### Longitudinal Registration

Diffusion data were processed using TORTOISE (Pierpaoli et al. 2010) for motion and eddy current correction. Within-subject longitudinal registration was performed using the T2-weighted image from the final visit as the structural reference. The T1-weighted volume from the final visit was processed using FreeSurfer (Fischl 2012) for tissue segmentation. Segmentation maps were transformed to native diffusion space using ANTs-based registration (Avants, Tustison, and Song 2009). Smaller anatomical regions were combined and morphologically eroded in 3D using a custom MATLAB script (MathWorks Inc., 2025) to generate eleven regions of interest (ROIs): Corpus callosum, Left cortical white matter, Right cortical white matter, Left cortical gray matter, Right cortical gray matter, Thalami, Putamen, Caudate, Amygdala, Hippocampi, Pallidum.

#### Diffusion Modeling

Diffusion tensor metrics (fractional anisotropy, mean diffusivity, axial diffusivity, radial diffusivity) were estimated first. MAP-MRI reconstruction was then performed to estimate the diffusion propagator and associated metrics, including: Propagator anisotropy (PA), Return-to-origin probability (RTOP), Return-to-axis probability (RTAP), Return-to-plane probability (RTPP), Non-Gaussianity (NG). MAP-MRI estimation at each visit was referenced to the diffusion tensor from the final visit to ensure longitudinal consistency.

### Longitudinal MAP-MRI Analysis

Qualitative review of sagittal 3D T2-FLAIR images was performed to identify imaging abnormalities indicative of underlying alterations in tissue by reviewers who were blinded to the quantitative longitudinal MAP-MRI findings.

All quantitative analyses were performed using custom scripts in Python and MATLAB. For each participant, voxel-wise absolute statistical distance between propagators at each visit (Visit_1_–E_3_) relative to Visit_4_ was computed using the Hellinger(Hadamard 1909) and total variation (Kelbert 2023) distances. Percent difference maps for each MAP-MRI metric were calculated relative to E_4_ and normalized by the mean value across all visits. Within each ROI, the median value of total variation distance and percent difference in MAP-MRI metrics were used as the participant-level summary statistic. This final subset included data from 111 participants (80 controls, 31 mTBI) with complete data across the four visits.

### Statistical Analysis

Clinical mTBI assessments: Preliminary evaluation of cross-sectional differences (Control vs mTBI) in the RPQ-3, RPQ-13, Total BESS-Firm and Total BESS-Foam assessments at the baseline visits (Visit_1_ and Visit_2_) was performed using independent sample t-testing with Bonferroni correction for multiple comparisons was applied to p-values.

Longitudinal effects of mTBI on diffusion metrics and clinical mTBI markers: The median values of each DTI and MAP-MRI metric within the eleven ROIs at all visits were computed for all participants. The RPQ-3, RPQ-13, Total BESS-Firm, Total BESS-Foam and Total BESS scores at each visit were used for correlation analyses and in generalized additive mixed (GAM) models (Wood 2017) with a cubic spline term for intensity and time by case status interaction. The control group at Visit_1_ is used as the reference category in the GAM models with the intercept corresponding the mean of that group. Effect estimates from these models for mTBI, Visit_2-4_, and interaction terms are changes or differnces from the scores of the reference group/visit. False discovery rate (FDR) was controlled within each ROI using the Benjamini–Hochberg procedure (Benjamini and Hochberg 1995).

Longitudinal effects of subtle alterations in tissue microstructure following injury on propagator distances was performed in the subset of participants who were evaluated across all four visits. The median values of the difference (MAP-MRI metrics) and the total variation within each ROI were used as the summary statistic for each participant in this subset. Linear mixed effects analysis with Parameters, Timepoint (Visit_1_vsVisit_4_; Visit_2_vsVisit_4_; Visit_3_vsVisit_4_), and ROI as fixed variables and Subject ID as the random variable followed by inference testing using analysis of variance was used to evaluate the relationship between these difference metrics and time within each ROI. The p-values were adjusted for multiple comparisons.

## Results

Table 1 summarizes the demographic characteristics of participants included in the analysis. The mTBI and control groups were similar in age and sex distribution (mTBI: 52.5% male, mean age 21.9 ± 8.5 years; controls: 42.9% male, mean age 21.9 ± 8.3 years). The majority of participants in both groups were younger than 20 years (60.7%). Among participants with mTBI, 65.0% reported no prior history of TBI, and the most common mechanism of injury was sports-related (56.2%).

**Table 1:**
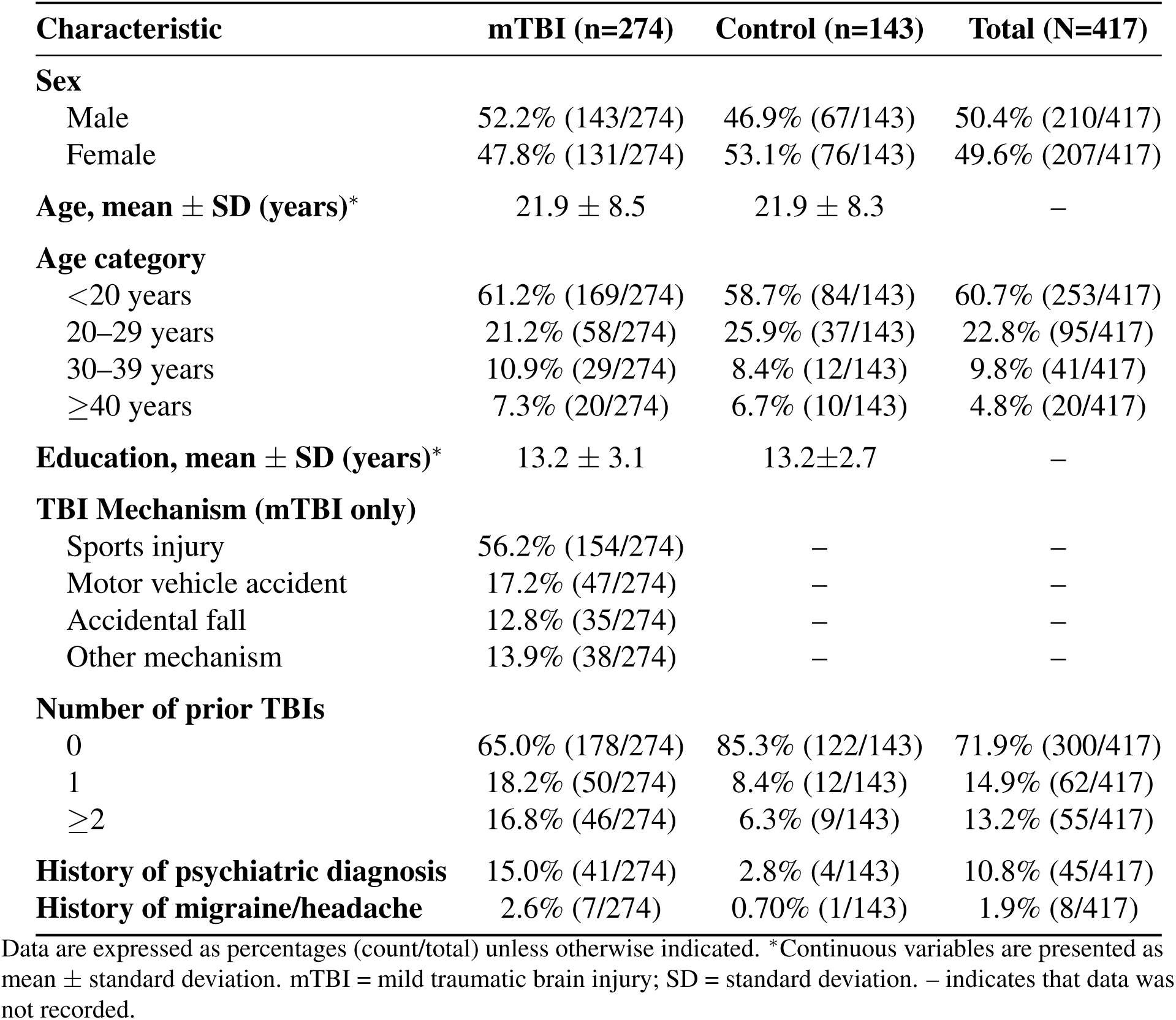
Participant Demographics for Longitudinal Quantitative Analysis of MAP-MRI and Clinical mTBI Markers (N = 417). Age and sex-matched mTBI patients and controls were recruited across multiple sites. For both populations, the majority of patients were adolescents (< 20 years old) and had no prior history of TBI prior to the injury that qualified patients to be enrolled into the study.

| Characteristic | mTBI (n=274) | Control (n=143) | Total (N=417) |
| --- | --- | --- | --- |
| **Sex** |  |  |  |
| Male | 52.2% (143/274) | 46.9% (67/143) | 50.4% (210/417) |
| Female | 47.8% (131/274) | 53.1% (76/143) | 49.6% (207/417) |
| **Age, mean ± SD (years)\*** | 21.9 ± 8.5 | 21.9 ± 8.3 | – |
| **Age category** |  |  |  |
| <20 years | 61.2% (169/274) | 58.7% (84/143) | 60.7% (253/417) |
| 20–29 years | 21.2% (58/274) | 25.9% (37/143) | 22.8% (95/417) |
| 30–39 years | 10.9% (29/274) | 8.4% (12/143) | 9.8% (41/417) |
| ≥40 years | 7.3% (20/274) | 6.7% (10/143) | 4.8% (20/417) |
| **Education, mean ± SD (years)\*** | 13.2 ± 3.1 | 13.2±2.7 | – |
| **TBI Mechanism (mTBI only)** |  |  |  |
| Sports injury | 56.2% (154/274) | – | – |
| Motor vehicle accident | 17.2% (47/274) | – | – |
| Accidental fall | 12.8% (35/274) | – | – |
| Other mechanism | 13.9% (38/274) | – | – |
| **Number of prior TBIs** |  |  |  |
| 0 | 65.0% (178/274) | 85.3% (122/143) | 71.9% (300/417) |
| 1 | 18.2% (50/274) | 8.4% (12/143) | 14.9% (62/417) |
| ≥2 | 16.8% (46/274) | 6.3% (9/143) | 13.2% (55/417) |
| **History of psychiatric diagnosis** | 15.0% (41/274) | 2.8% (4/143) | 10.8% (45/417) |
| **History of migraine/headache** | 2.6% (7/274) | 0.70% (1/143) | 1.9% (8/417) |

Data are expressed as percentages (count/total) unless otherwise indicated. \*Continuous variables are presented as mean ± standard deviation. mTBI = mild traumatic brain injury; SD = standard deviation. – indicates that data was not recorded.

**Table 2:**
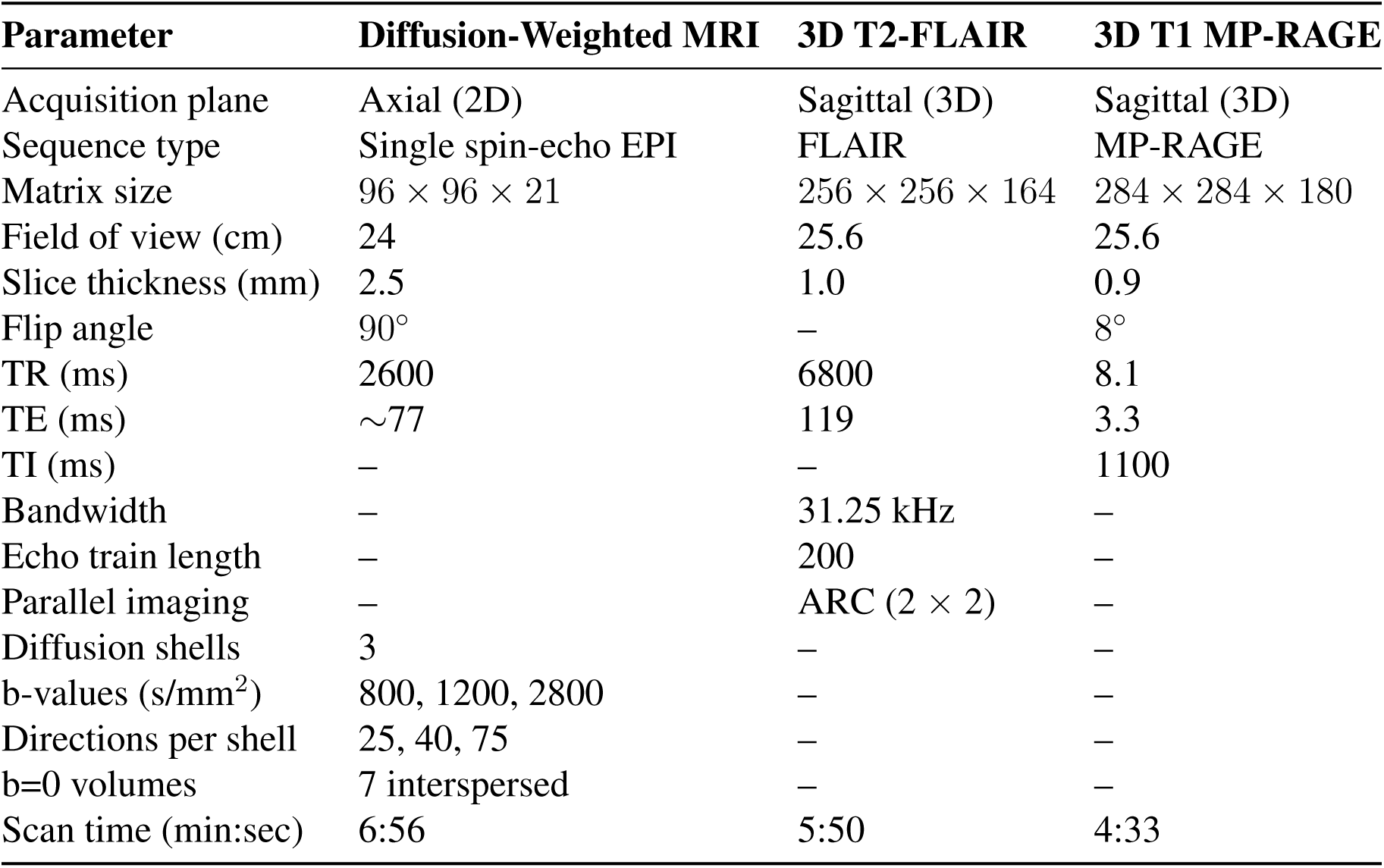
MRI Acquisition Parameters.

Demographic characteristics for the subset of participants with complete data across all four visits are provided in Supplementary Table 1. This subset remained comparable between groups, with similar age and sex distributions; 59.5% of participants were younger than 20 years, and 71.0% of mTBI participants had no prior history of TBI.

Figure 2 illustrates representative diffusion tensor and propagator-derived metrics from a control participant at Visit_1_. After quality control and completeness filtering, the final analytic sample comprised 417 participants (274 mTBI, 143 controls). The processing pipeline yielded diffusion tensor imaging (DTI) and MAP-MRI metrics with minimal artifacts across visits. Qualitatively, fractional anisotropy (FA) demonstrated reduced values in regions of complex fiber architecture, such as cortical white matter, whereas propagator-derived metrics (e.g., propagator anisotropy [PA]) appeared more spatially homogeneous in these regions.

**Figure 2:**
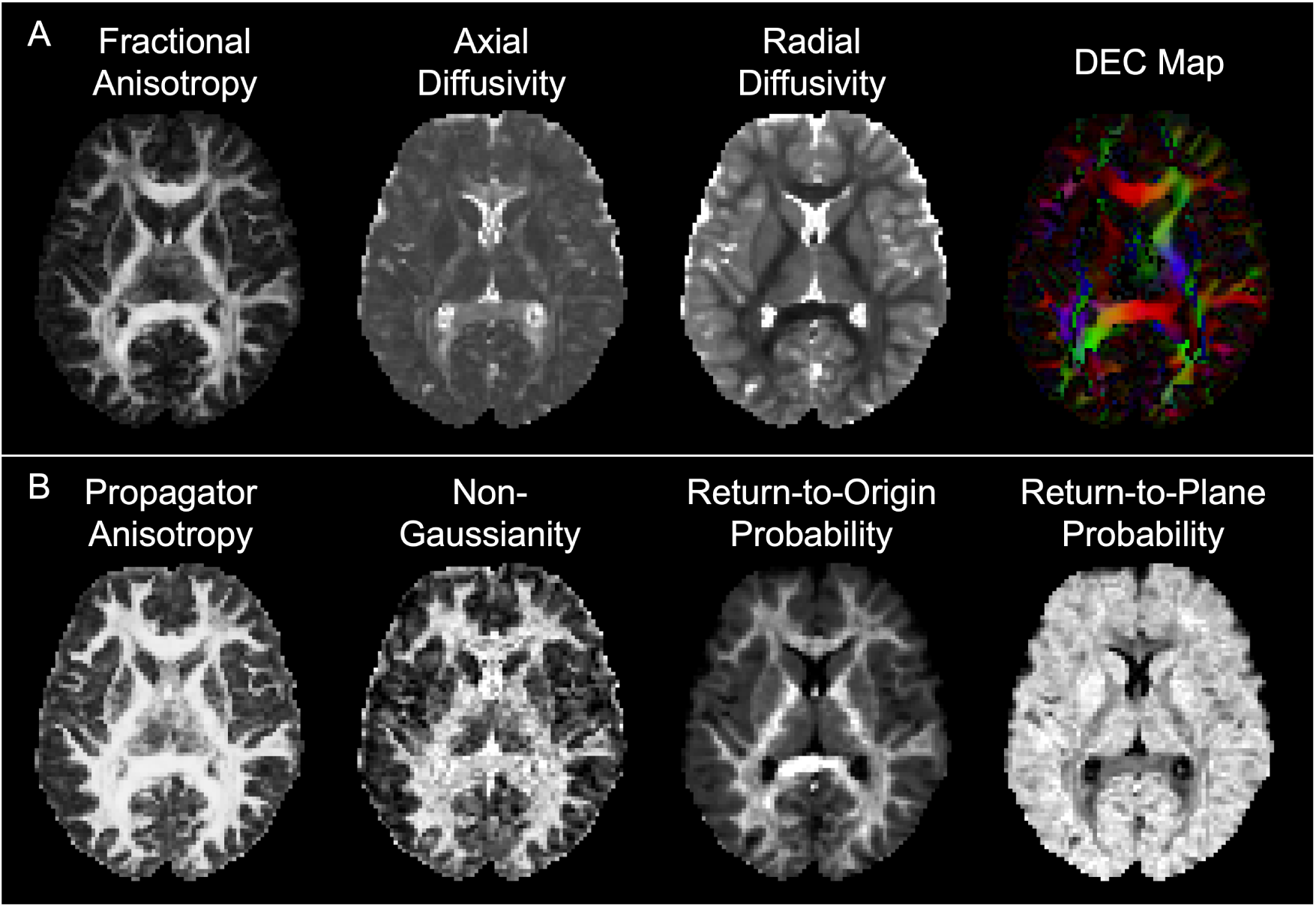
Exemplar diffusion tensor and MAP-MRI metrics computed in an exemplar control. A. Diffusion tensor metrics included fractional anisotropy, axial diffusivity, radial diffusivity and directional encoded color (DEC) maps. B. MAP-MRI metrics included propagator anisotropy, Non-Gaussianity, return-to-origin probability and return-to-plane probability.

Figure 3A summarizes diffusion MRI parameters associated with postural stability, as assessed using generalized additive mixed models accounting for repeated measures, case status, and time within each ROI. BESS scores on both foam and firm surfaces were significantly associated (p_adj_ *<*0.05) with tensor-derived mean and axial diffusivities, as well as MAP-MRI metrics including RTPP, non-Gaussianity (NG), and propagator anisotropy (PA) within the caudate. Similar associations were observed in the putamen and cortical gray matter, where BESS scores were significantly related to RTAP, RTOP, RTPP, PA, and NG.

**Figure 3:**
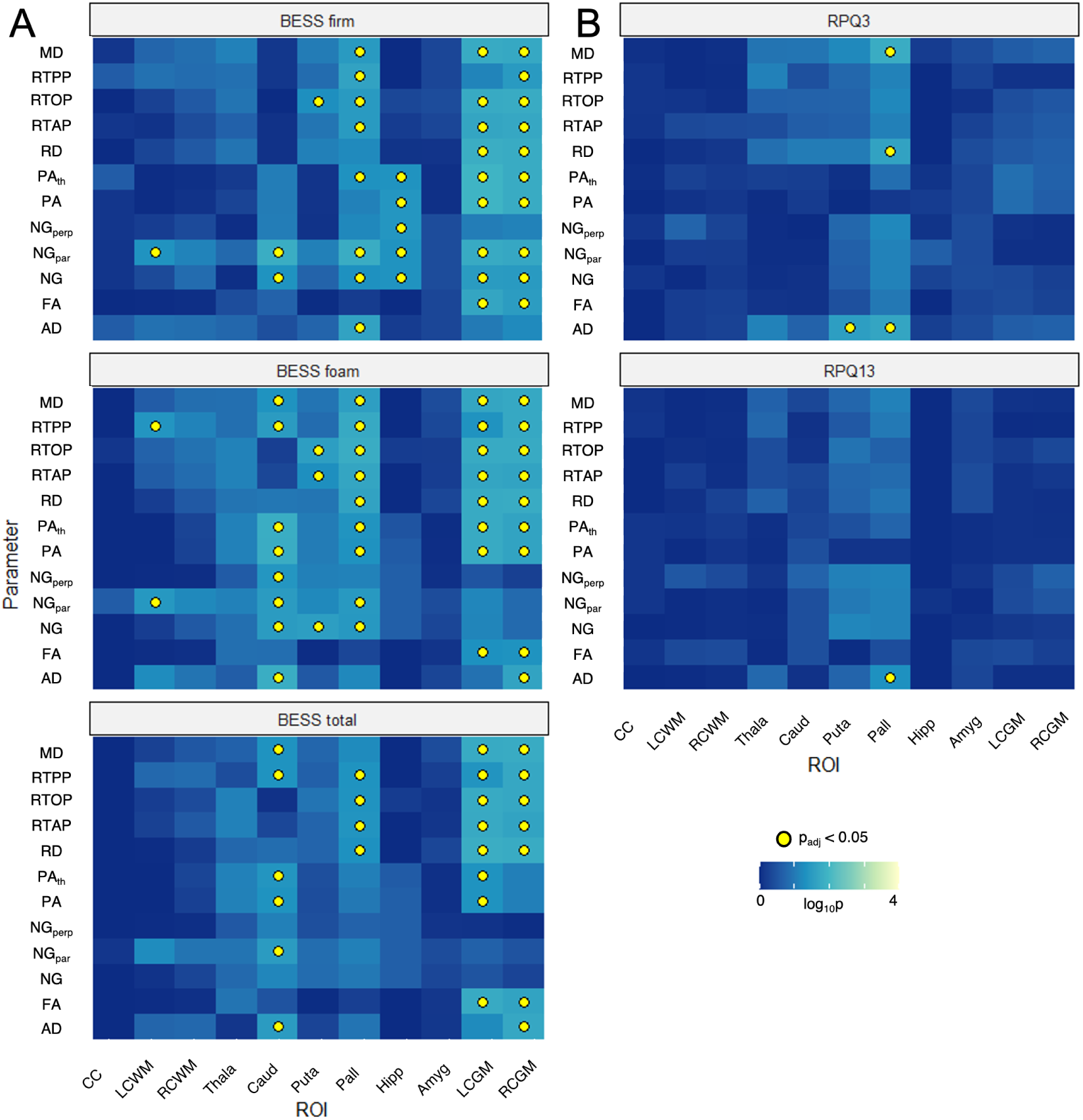
Longitudinal cubic spline analysis of diffusion MRI metrics in regions of interest with clinical mTBI markers shows association of MAP-MRI metrics in cortical gray matter and deep gray matter nuclei with clinical mTBI markers. A. Postural stability was assessed using the Balance Error Scoring System (BESS) on firm and foam surfaces. DTI and MAP-MRI parameters in the caudate, pallidum and cortical gray matter regions of interest (ROIs) show significant correspondence with BESS scores (firm, foam and total). B. DTI parameters in the putamen and caudate show significant correspondence with self reported Rivermead post-concussive questionnaire of physical symptoms (RPQ-3) and psychosocial lifestyle functioning and lifestyle symptoms (RPQ-13). Abbreviations: MD - mean diffusivity; RTOP - return-to-origin probability; RTPP - return-to-plane probability; RTAP - return-to-axis probability; RD - radial diffusivity; PA_th_ - nonlinear transformed propagator anisotropy; PA - propagator anisotropy; NG - Non-Gaussianity; NG_par_ - parallel component of Non-Gaussianity; NG_perp_ - perpendicular component of Non-Gaussianity; FA - fractional anisotropy; AD - axial diffusivity; CC - corpus callosum; LCWM - left cortical white matter; RCWM - right cortical white matter; Thala- thalamus; Caud - Caudate; Puta - Putamen; Pall - Pallidum; Hipp - Hippocampus; Amyg - Amygdala; LCGM - left cortical gray matter; RCGM - right cortical gray matter.

Figure 3B shows associations between diffusion metrics and self-reported post-concussive symptoms. Physical symptom scores (RPQ-3) were significantly associated with tensor-derived mean, axial, and radial diffusivities in the putamen and pallidum. Psychosocial symptom scores (RPQ-13) were significantly associated with axial diffusivity in the pallidum. No significant associations were observed between MAP-MRI metrics and self-reported symptom scores across any ROI.

Baseline clinical measures (Visit_1_ or Visit_2_) are summarized in Supplementary Table 2. At baseline, mTBI participants demonstrated significantly higher RPQ-3 and RPQ-13 scores and worse total BESS performance on the firm surface compared with controls, with all differences remaining significant after correction for multiple comparisons (p_adj_ ¡ 0.05).

Table 3 summarizes the fixed effects from generalized additive mixed models evaluating associations between clinical scores, case status, and time. Distinct patterns were observed across outcome measures.

**Table 3:** Generalized additive mixed model analysis of clinical mTBI markers over time. Scoring of postural stability in mTBI and controls changes minimally over time, while post-concussion retrospective reporting of physical symptoms and psychosocial functioning are increased in mTBI patients relative to controls acutely after injury and approach scores of controls by 90 days post injury.

|  |  | Intercept | mTBI | Visit 2 | Visit 3 | Visit 4 | mTBI:<br>Visit 2 | mTBI:<br>Visit 3 | mTBI:<br>Visit 4 |
| --- | --- | --- | --- | --- | --- | --- | --- | --- | --- |
| **Total BESS - Firm** | Estimate | 11.66 | 2.68 | -1.30 | -2.04 | -1.29 | 1.08 | 1.30 | -0.50 |
|  | p | <1x10<sup>-5</sup> | 0.08 | 0.26 | 0.08 | 0.36 | 0.50 | 0.41 | 0.78 |
| **Total BESS - Foam** | Estimate | 4.50 | 0.74 | -0.70 | -0.86 | -1.20 | 0.32 | 0.28 | 0.15 |
|  | p | <1x10<sup>-5</sup> | 0.17 | 0.048 | **0.02** | **0.003** | 0.57 | 0.61 | 0.79 |
| **Total BESS** | Estimate | 8.95 | 0.58 | -0.97 | -1.51 | -1.33 | 1.36 | 1.35 | 0.49 |
|  | p | <1x10<sup>-5</sup> | 0.55 | 0.19 | **0.04** | 0.15 | 0.18 | 0.18 | 0.66 |
| **RPQ-3** | Estimate | 0.69 | 4.07 | -0.18 | -0.46 | 0.01 | -0.80 | -2.00 | -3.55 |
|  | p | 0.0004 | <1x10<sup>-5</sup> | 0.39 | **0.03** | 0.92 | **0.01** | <1x10<sup>-5</sup> | <1x10<sup>-5</sup> |
| **RPQ-13** | Estimate | 2.44 | 12.76 | -0.14 | -1.19 | -0.45 | -1.57 | -5.36 | -10.20 |
|  | p | 0.0007 | <1x10<sup>-5</sup> | 0.85 | 0.12 | 0.54 | 0.16 | <1x10<sup>-5</sup> | <1x10<sup>-5</sup> |

The intercept represents baseline control values, the mTBI term reflects baseline group differences, visit terms capture longitudinal changes in controls, and interaction terms quantify whether temporal trajectories differ between mTBI and control groups. Intercept terms represent baseline values in the control group and are not interpreted inferentially. P-values shown in bold indicate significant fixed terms used for inference.

For BESS assessments, baseline differences between mTBI and control participants were small and not statistically significant (marginally for “Firm”), consistent with the modest mTBI main effects. Scores showed minimal change over time in both groups, with slight decreases after Visit_1_. Interaction terms between mTBI and visit were not statistically significant, indicating similar longitudinal trajectories across groups.

In contrast, RPQ-3 and RPQ-13 scores demonstrated substantial baseline differences with higher scores in the mTBI group relative to controls, 4.07 and 12.76 points on average, respectively. Over time, RPQ scores remained relatively stable in controls seen in the near zero effects of Visit_2-4_ but decreased in the mTBI group, negative and significant effects (p_adj_*<*0.05) of mTBI:Visit_2-4_. The decrease over time in mTBI group resulted in a near convergence of the scores between the two groups by Visit_4_ shown by the mTBI effect, which corresponds to Visit_1_, negated by the mTBI:Visit_4_ effect.

Supplementary Table 3 presents correlations between diffusion metrics and clinical scores in control participants. After correction for multiple comparisons, significant associations (p_adj_ *<*0.05) were primarily observed at Visit_1_, demonstrating weak correlations between both MAP-MRI and DTI metrics in cortical white matter and the thalamus with RPQ-3 scores.

In mTBI participants (Supplementary Table 4), significant correlations were more frequently observed at Visit_2_ and Visit_3_. These associations reflected weak correlations between diffusion metrics (MAP-MRI and DTI) in the thalamus and total BESS scores on the foam surface.

Figure 4 illustrates fiber orientation distribution (FOD) functions in cortical gray and white matter overlaid on propagator anisotropy (PA) maps in an exemplar mTBI participant in their twenties who completed all four post-injury visits. The FODs enable visualization of subtle changes in the orientational structure of the three-dimensional displacement distributions (diffusion propagators) across time points. The Hellinger distance between propagators provides a comprehensive statistical measure of voxelwise subtle temporal changes that are not captured by any single MAP-MRI or DTI-derived metric. In this exemplar, propagator distance maps highlight a white matter region exhibiting the largest deviations at earlier time points (Visit_1_ and Visit_2_) relative to Visit_4_.

**Figure 4:**
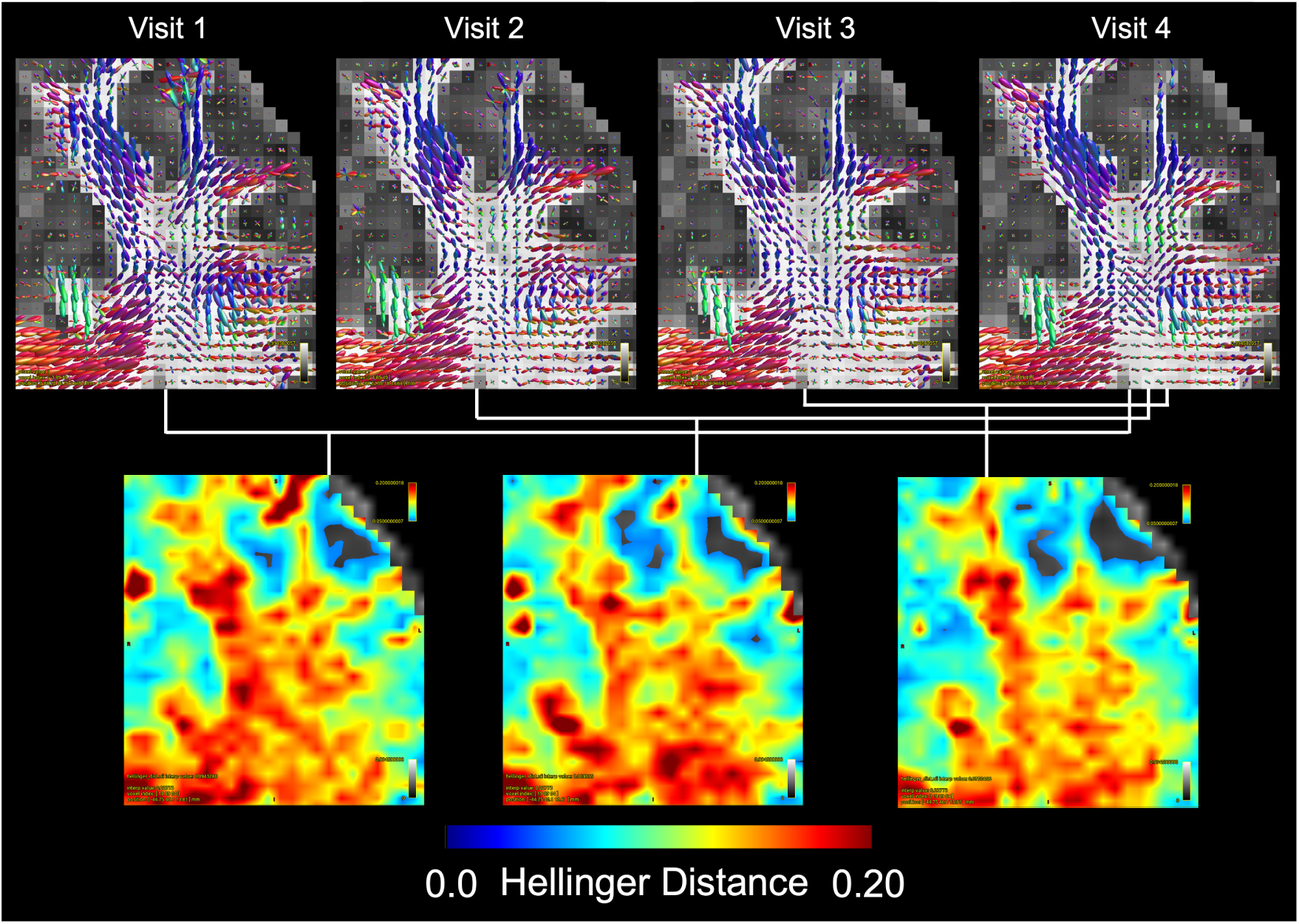
Fiber orientation distribution (FOD) functions derived from MAP-MRI data enable visualization of diffusion propagator anisotropy and provide a more comprehensive characterization of changes in fiber architectures beyond those captured by individual parameters such as PA, RTAP, or RTPP. FODs overlaid on coronal slices of PA in the left prefrontal cortex of a male mTBI subject in their twenties across the 4 post-injury visits. In each voxel, the propagator distance computed between each of the first 3 visits and the 4th (baseline) visit (90 days post injury) captures general differences between propagators, including changes in FODs. Larger propagator distance values are seen in WM at earlier timepoints.

Qualitative assessment across participants (Supplementary Figure 2) showed no consistent differences in propagator distance, between mTBI and control groups or across time points in gray matter and major white matter tracts. Consistent with these observations, linear mixed-effects analysis in the subset of participants with complete longitudinal data demonstrated no significant association between propagator distance metrics and time interval across any of the eleven ROIs.

Figure 5 presents a detailed qualitative assessment of longitudinally registered data from the same exemplar participant. White matter hyperintensities (WMHs) were visible on T2-weighted FLAIR images and appeared as hypointense foci on T1-weighted MP-RAGE images. These regions exhibited heterogeneous signal characteristics on diffusion maps, appearing hypointense on RTOP and direction-encoded color maps and mildly hyperintense on mean diffusivity maps. While no focal changes were observed on PA maps within these regions, reduced PA signal was noted in surrounding tissue. Additionally, RTOP maps revealed hypointense foci not evident on structural MRI.

**Figure 5:**
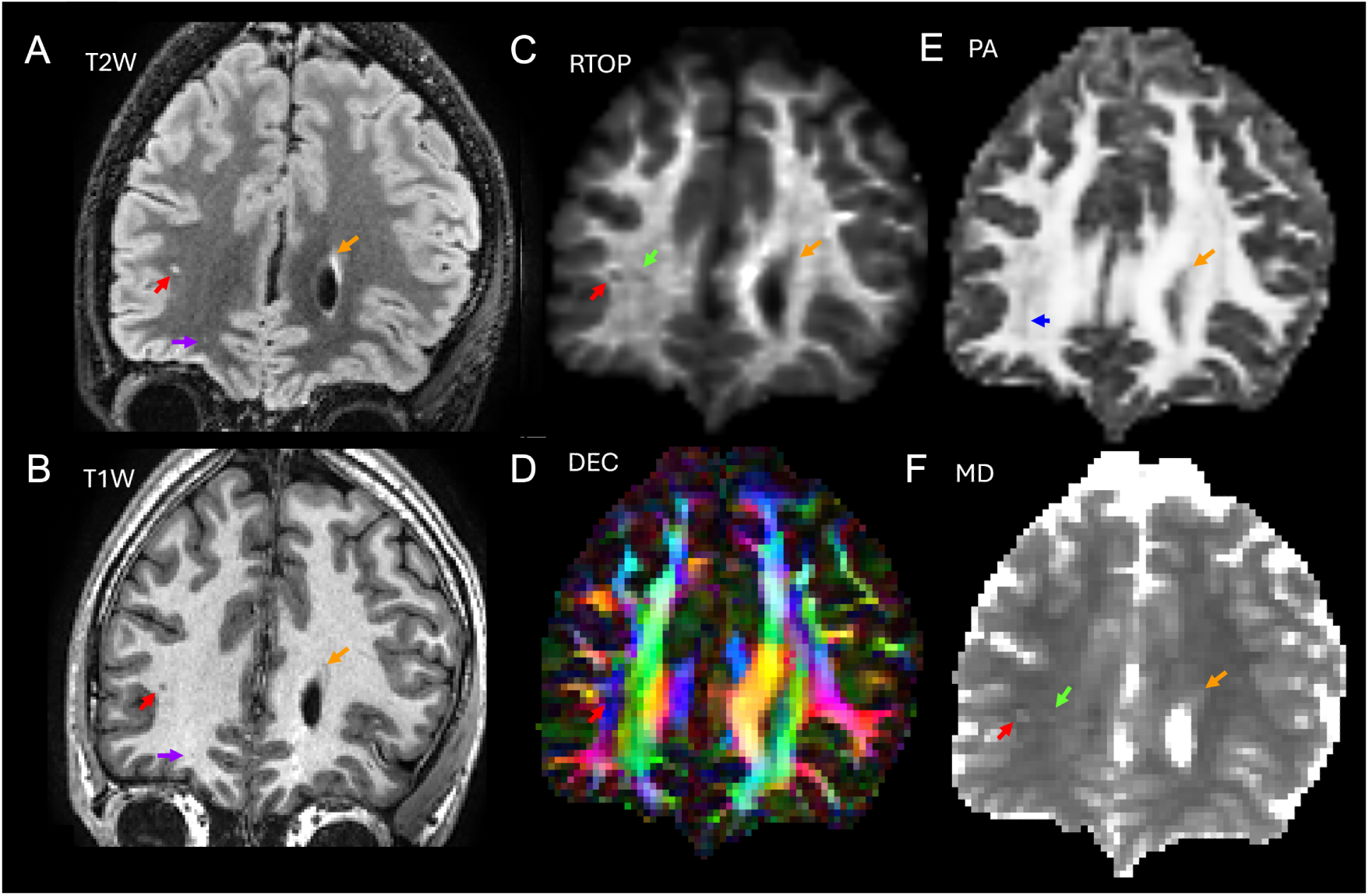
Characterization of white matter abnormalities suggests that MAP-MRI provides complementary information to conventional radiological assessment. A. Multiple white matter hyperintensities (WHM) in the left frontal lobe are visible on the coronal slices of the FLAIR-T2W scan from a male mTBI subject in their twenties scan (red, orange, and purple arrows). B. These abnormalities are also observed in the T1W-MPRAGE (red, orange, and purple arrows). C. Return-to-origin probability (RTOP) in the location corresponding to the larger WMH (red arrows) is hypointense, along with an adjacent hypointensity (green arrows) not visible on the structural scans. D. The same WMH region (red arrow) is hypointense on the DEC map. E. The region surrounding the WMH is hypointense on the propagator anisotropy (PA) map (blue arrow), suggesting potential loss of microstructural integrity. F. The larger WMH (red arrow) corresponds to localized hyperintense mean diffusivity (MD).

## Discussion

Our longitudinal MAP-MRI processing and registration pipeline enables efficient characterization of changes in the diffusion propagator and derived metrics in mTBI, a condition marked by subtle and spatially heterogeneous injury. MAP-MRI provides a comprehensive framework that encompasses conventional DTI while yielding reproducible metrics sensitive to underlying microstructural features, including cellular size, shape, and orientation in both gray and white matter.

This study leverages several strengths of the dataset, including diffusion MRI acquisitions with relatively high diffusion weightings and a large cohort of age- and sex-matched mTBI and control participants with limited comorbidities that could confound imaging findings. In addition, approximately 25% of participants were scanned at four post-injury time points, enabling detailed evaluation of longitudinal trajectories in both MAP-MRI metrics and the diffusion propagator.

The longitudinal processing framework produced consistently registered DTI and MAP-MRI measures across visits. Propagator-derived metrics, such as propagator anisotropy (PA), demonstrated reduced variability in regions with complex fiber architecture compared to DTI-derived fractional anisotropy (FA), while displacement-based measures, including RTOP and RTPP, provided complementary microstructural information beyond tensor-derived metrics such as axial and radial diffusivity.

### Correlation of imaging metrics and clinical scores

Mixed-effects modeling of longitudinal changes in diffusion MRI metrics and clinical measures showed distinct patterns across outcome types. Objective assessments of postural stability (BESS) did not differ significantly between mTBI and control groups and demonstrated minimal change over time. In contrast, RPQ symptom scores exhibited marked group differences at early time points that diminished over time, with scores converging by Visit_4_.

The discrepancy between RPQ and BESS findings may reflect the subjective nature and temporal framing of symptom reporting in the RPQ, particularly in individuals with mTBI, for whom symptom onset is anchored to a known injury event. In contrast, BESS provides an objective measure of postural stability obtained within the same visit as the MRI examination. This difference in temporal alignment may contribute to the observed results, as BESS captures physiological function at the time of imaging, whereas RPQ scores reflect retrospectively reported symptoms that may not correspond directly to current microstructural states. Consistent with this distinction, diffusion MRI metrics—particularly MAP-MRI parameters including RTOP, RTAP, RTPP, propagator anisotropy (PA), and non-Gaussianity (NG)—showed significant associations with BESS performance, suggesting sensitivity to contemporaneous physiological changes.

By comparison, no consistent associations were observed between diffusion metrics and RPQ scores, indicating that regionally averaged diffusion measures may be less sensitive to retrospectively reported symptoms. Notably, associations between MAP-MRI metrics and BESS performance were observed in the caudate, pallidum, and cortical gray matter. These findings are consistent with prior longitudinal MAP-MRI work demonstrating that intersession variability in these regions following mTBI exceeds test–retest variability in controls (Gangolli et al. 2025).

### Detection of subtle microstructural changes

Our analysis pipeline incorporates within-subject registration, enabling accurate quantitation of local changes in the diffusion propagators and associated metrics, which can subsequently be summarized at the ROI level. This approach was designed to eliminate intersubject variability and maximize sensitivity to subtle mTBI-related longitudinal microstructural changes in each voxel. Quantitative analyses revealed no consistent spatiotemporal patterns of change in the diffusion propagator or MAP-MRI metrics across the four post-injury visits that exceeded the variability observed in healthy controls. Overall, MAP-MRI metrics remained relatively stable over time in both mTBI and control groups.

Voxel-wise distance between diffusion propagators was greatest in major white matter tracts in both groups, likely reflecting sensitivity to anisotropic microstructure and fiber orientation, consistent with the small percent changes observed (approximately ±5%). Additional regions exhibiting increased variability were primarily located near cerebrospinal fluid spaces and are likely influenced by physiological noise sources, such as cardiac pulsation.

### White matter heterogeneity

Importantly, white matter hyperintensities (WMHs) were observed in this cohort, consistent with prior findings in the same study population (Tanwar et al. 2025). That prior work found that T2-FLAIR white matter abnormalities were not specific to mTBI. In the present study, qualitative assessment of a longitudinally registered case revealed hypointense foci on RTOP and propagator anisotropy (PA) maps in regions that appeared normal on both T2-FLAIR and T1-weighted imaging.

Although the spatial extent of these focal MAP-MRI abnormalities is constrained by voxel resolution, these findings suggest that propagator-derived metrics may provide complementary microstructural information beyond conventional structural imaging (Figure 5). These alterations are not necessarily specific to mTBI and may reflect underlying neuropathological processes independent of injury status. Alternatively, regions demonstrating MAP-MRI abnormalities in the absence of structural changes may indicate increased sensitivity of MAP-MRI to subtle tissue remodeling, suggesting potential utility as an imaging marker of microstructural recovery following neurological insult.

### Limitations

A primary limitation of this study relates to the selection criteria and demographic characteristics of the study population. Participants with mTBI were required to have sustained a single, mild injury within 10 days of enrollment, and the majority had no prior history of TBI. Preclinical models suggest that while a single mild injury may produce acute pathophysiological changes, these effects often do not persist over weeks to months (Kulkarni et al. 2019; Szarka et al. 2019). In contrast, repeated mTBI has been associated with cumulative microstructural injury and more sustained alterations.

In addition, the study cohort was predominantly composed of younger individuals, with most participants under 20 years of age. The developing brain may exhibit greater resilience and recovery following injury, potentially reducing the magnitude and persistence of detectable microstructural changes. Although mTBI is common in younger populations, older adults (*>*40 years) are at increased risk of long-term disability and functional decline after TBI (Corrigan, Selassie, and Orman 2008).

Together, these factors suggest that the injuries captured in this study may have been associated with relatively subtle or transient microstructural alterations. Future studies including populations with repeated mTBI exposure or broader age distributions may provide greater sensitivity for detecting persistent neuropathological changes using MAP-MRI.

A second limitation relates to the use of region-of-interest (ROI)–based summaries to quantify longitudinal changes in MAP-MRI metrics and propagator distances. Although the processing pipeline enables voxel-wise computation of changes, aggregation within relatively large ROIs may reduce sensitivity to localized alterations that affect only a subset of voxels. As a result, subtle or spatially heterogeneous microstructural changes may be attenuated in ROI-averaged measures.

In addition, voxel-wise percent changes in MAP-MRI metrics and propagator distances were computed relative to Visit_4_, rather than to a true pre-injury baseline. If microstructural alterations had partially or fully resolved by this time point—particularly in participants without prior TBI—this approach may underestimate the magnitude of injury-related changes. A prospective study design including pre-injury baseline imaging would provide a more appropriate reference for longitudinal comparisons.

This limitation is further illustrated by the qualitative findings of focal abnormalities in MAP-MRI metrics corresponding to T2-FLAIR white matter hyperintensities, which were not detected in the ROI-based analysis. These observations underscore the importance of voxel-level or spatially adaptive approaches for capturing focal microstructural alterations.

Finally, although the diffusion propagator was estimated in this dataset, the acquisition protocol was not optimized for MAP-MRI. In particular, the use of three shells with a maximum diffusion weighting of 2800 s/mm^2^ is sufficient for stable estimation of DTI and kurtosis metrics but may limit the robustness of higher-order models such as MAP-MRI. Prior work has shown that propagator-derived measures such as RTOP, RTAP, and RTPP are relatively stable at moderate b-values, whereas their distributions—and the sensitivity of metrics such as non-Gaussianity (NG) and propagator anisotropy (PA)—depend more strongly on the inclusion of higher diffusion weightings (Hutchinson, Avram, Irfanoglu, et al. 2017).

Accordingly, multi-site and longitudinal studies aiming to leverage MAP-MRI would benefit from standardized acquisition protocols incorporating higher b-values to improve the sensitivity and reproducibility of propagator-derived metrics and to facilitate quantitative comparisons across sites and time points.

Future work will focus on applying this longitudinal MAP-MRI framework to datasets with acquisition protocols optimized for propagator estimation and to populations in which injury-related sequelae are more pronounced. In addition, longitudinally registered diffusion metrics may enable development of data-driven approaches, such as convolutional neural networks and AI-based techniques, for direct image-based differentiation between mTBI and control subjects.

Our findings also support the use of prospective longitudinal study designs in populations at elevated risk for mTBI, such as military cohorts, where baseline imaging can be acquired prior to exposure. Such designs would provide a critical reference for quantifying injury-related changes and may improve the sensitivity of imaging biomarkers for detecting and monitoring microstructural alterations following mTBI.

## Conclusions

We developed a quantitative, longitudinal image processing pipeline for diffusion MRI with sensitivity to high–b-value data and propagator-based metrics. Application of this framework to a cohort with mild and clinically subtle brain injury demonstrated no consistent group-level microstructural alterations detectable with diffusion MRI. These findings likely reflect the limited magnitude and transient nature of injury-related changes in this population.

Despite these results, the proposed pipeline provides a sensitive and reproducible framework for longitudinal assessment of tissue microstructure. Its application may be particularly valuable in settings where microstructural alterations are more pronounced, such as repeated mTBI or blast-related injury, where prior animal and postmortem studies have demonstrated measurable neuropathological changes (Fadon-Padilla et al. 2025; Gangolli et al. 2019; Echlin, Rahimi, and Wojtowicz 2021; Goodrich et al. 2016; Shively et al. 2016).

## Data Availability

All data produced in the present study are available upon reasonable request to the authors

## Acknowledgements

This work was supported by the Intramural Research Program (IRP) of the *Eunice Kennedy Shriver* National Institute of Child Health and Human Development (Award ZIA-HD008970-08), and partially funded by the Military Traumatic Brain Injury Initiative (MTBI^2^) through the Uniformed Services University of the Health Sciences (USU), Bethesda, MD (Award 309698-4.01-65310).

This work utilized the computational resources of the NIH HPC Biowulf cluster (https://hpc.nih.gov/).

Portions of this manuscript were edited for clarity and language using an artificial intelligence–based tool (ChatGPT 5.3). The tool was not used for data analysis or interpretation. The authors reviewed and take full responsibility for the content.

## Disclaimer

The authors have no conflicts of interest to disclose. The views, information or content, and conclusions presented do not necessarily represent the official position or policy of, nor should any official endorsement be inferred on the part of, the Uniformed Services University, the Department of War, the U.S. Government or The Henry M. Jackson Foundation for the Advancement of Military Medicine, Inc.

## Notes

### Competing Interest Statement

The authors have declared no competing interest.

### Clinical Trial

NCT02556177

### Author Declarations

This was a data analysis of imaging data acquired as part of the Advanced MRI Applications for Mild Traumatic Brain Injury-Phase 2 (mTBI-phase2) sponsored by GE Healthcare. The Institutional Review Boards (IRB) of: University of California, San Francisco University of California, San Diego Medical College of Wisconsin Hospital for Special Surgery University of Miami Health System University of Pittsburgh Medical College Houston Methodist Neurological Institute gave ethical approval for this work.

