## Supplementary for "Longitudinal MAP-MRI-based Assessment of Tissue Microstructural Alterations in Acute mTBI"

### Supplementary Material

Supplementary Table 1: Participant Demographics for Longitudinal Quantitative Analysis (N = 111)

| Characteristic | mTBI (n=31) | Control (n=80) | Total (N=111) |
|---|---|---|---|
| **Sex** | | | |
| Male | 51.6% (16/31) | 45.0% (36/80) | 46.8% (52/111) |
| Female | 48.4% (15/31) | 55.0% (44/80) | 53.2% (59/111) |
| **Age, mean ± SD (years)*** | 23.2 ± 8.4 | 21.7 ± 8.4 | – |
| **Age category** | | | |
| <20 years | 51.6% (16/31) | 62.5% (50/80) | 59.5% (66/111) |
| 20–29 years | 32.2% (10/31) | 22.5% (18/80) | 25.2% (28/111) |
| 30–39 years | 9.7% (3/31) | 8.8% (7/80) | 9.0% (10/111) |
| ≥40 years | 6.5% (2/31) | 6.3% (5/80) | 6.3% (7/111) |
| **Education, mean ± SD (years)*** | 14.1 ± 2.6 | 13.0 ± 2.7 | – |
| **TBI Mechanism (mTBI only)** | | | |
| Sports injury | 38.7% (12/31) | – | – |
| Motor vehicle accident | 12.9% (4/31) | – | – |
| Accidental fall | 29.0% (9/31) | – | – |
| Other mechanism | 19.3% (6/31) | – | – |
| **Number of prior TBIs** | | | |
| 0 | 71.0% (22/31) | 83.8% (67/80) | 80.2% (89/111) |
| 1 | 9.7% (3/31) | 10.0% (8/80) | 9.9% (11/111) |
| ≥2 | 19.3% (6/31) | 6.4% (5/80) | 9.9% (11/111) |
| **History of psychiatric diagnosis** | 16.1% (5/31) | 1.3% (1/80) | 5.4% (6/111) |
| **History of migraine/headache** | 3.2% (1/31) | 1.3% (1/80) | 1.8% (2/111) |

Data are expressed as percentage (count/total) unless otherwise indicated. *Continuous variables are presented as mean ± standard deviation. mTBI = mild traumatic brain injury; SD = standard deviation. – indicates that data was not recorded.

Supplementary Table 2: Clinical Markers Measured at the first visit in mTBI patients and controls.

| Measure | mTBI (n = 274) | Controls (n = 143) | $p_{adj}$ |
|---|---|---|---|
| **BESS total score - Firm surface** | 6.2 ± 4.9 (0-25.0) | 4.4 ± 3.9 (0-18.0) | 0.015 |
| **BESS total score - Foam surface** | 9.0 ± 4.9 (0 - 28.0) | 9.1 ± 4.4 (3-20.0) | 4.0 |
| **RPQ-3** | 5.2 ± 2.2 (0-9.0) | 0.6 ± 1.0 (0-4.0) | $3.40 \times 10^{-35}$ |
| **RPQ-13** | 16.9 ± 9.8 (0-39.0) | 2.1 ± 3.6 (0-21.0) | $1.20 \times 10^{-24}$ |

Scores are expressed as the mean± standard deviation with the range in parentheses.

Supplementary Table 3: Correspondence of RPQ-3 score with regional diffusion MRI metrics in control participants.

| n = 143 controls | **Visit 1** | **Visit 2** |
|---|---|---|
| **Left Cortical White Matter** | NG ($\rho$ = -0.37, $p_{adj}$ = 0.025) | – |
| | RTOP ($\rho$ = -0.34,$p_{adj}$ = 0.034) | – |
| **Thalamus** | NG ($\rho$ = -0.29,$p_{adj}$ = 0.043) | – |
| | – | FA ($\rho$ = -0.32,$p_{adj}$ = 0.043) |
| | PA ($\rho$ = -0.31,$p_{adj}$ = 0.043) | – |

A correlation of clinical mTBI markers and diffusion MRI metrics across all subjects showed correspondence of MAP-MRI and DTI metrics with the RPQ-3 score during the initial visits in control participants. Significance values that ar shown were adjusted for multiple comparisons. Abbreviations: NG - Non-Gaussianity; RTOP - return-to-origin probability; FA - fractional anisotropy; PA - propagator anisotropy.

Supplementary Table 4: Correspondence of scores during the Total Balance Error Scoring System (BESS) assessment on a foam surface with diffusion MRI metrics in thalamic regions on interest in mTBI participants.

| n = 282 mTBI | **Visit 2** | **Visit 3** |
|---|---|---|
| **Thalamus** | RTOP ($\rho$ = 0.34, $p_{adj}$ = 0.037) | PA ($\rho$ = 0.36, $p_{adj}$ = 0.037) |
| | MD ($\rho$= -0.32, $p_{adj}$ = 0.037) | RTAP ($\rho$ = 0.32, $p_{adj}$ = 0.043) |
| | RTAP ($\rho$ = 0.30, $p_{adj}$ = 0.043) | RTOP (rho = 0.33, $p_{adj}$ = 0.043) |
| | PA ($\rho$ = 0.28, $p_{adj}$ = 0.044) | |

A correlation of clinical mTBI markers and diffusion MRI metrics in all subjects showed correspondence in thalamic ROIs of mTBI subjects between MAP-MRI and DTI metrics and the BESS scores (total foam and total overall) during the second and third visits. Significance values were adjusted for multiple comparisons. Abbreviations:PA - propagator anisotropy; RTOP - return-to-origin probability; RTAP - return-to-axis probability; FA - fractional anisotropy; MD - mean diffusivity; RD - radial diffusivity.

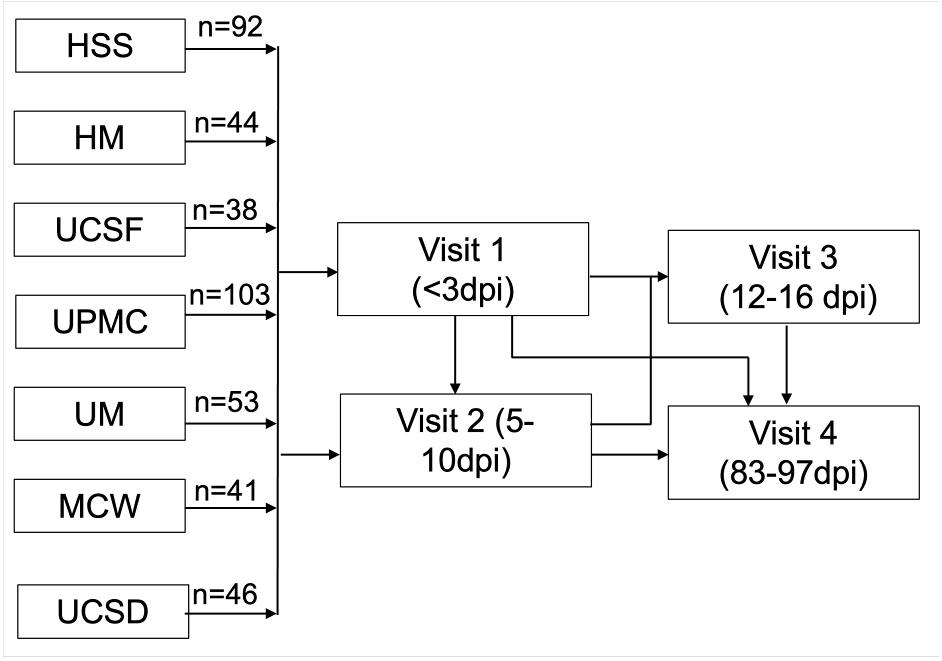


Supplementary Figure 1: Longitudinal MAP-MRI acquisition timeline. MRI data from control and mTBI subjects were acquired across six sites at four post-injury timepoints. Abbreviations: HSS - Hospital for Special Surgery; HM - Houston Methodist; UCSF - University of California, San Francisco; UPMC - University of Pittsburgh Medical Center; UM - University of Miami; MCW - Medical College of Wisconsin; UCSD - University of California, San Diego; dpi - days post injury.

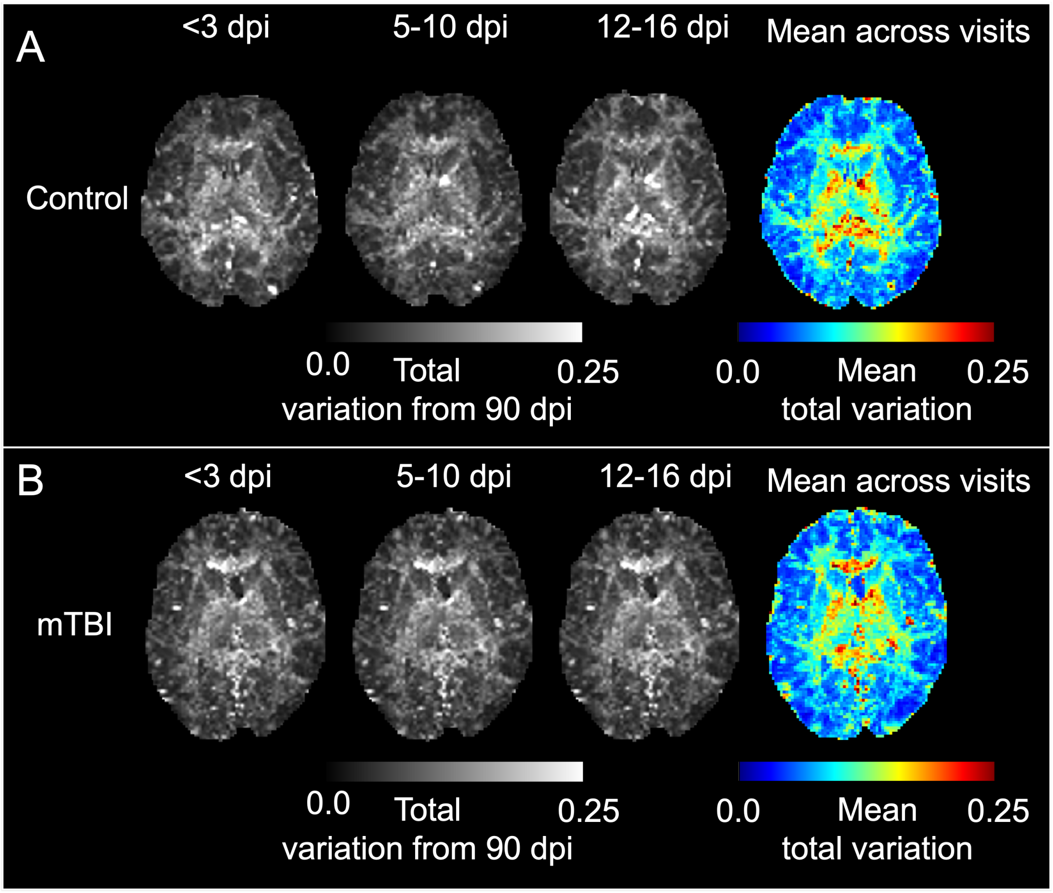


Supplementary Figure 2: Voxel-based total variation between propagators at post injury timepoints relative to 90 days post injury. A. Total variation and the mean total variation across visits computed in an exemplar control. B. Total variation and the mean total variation across visits computed in an exemplar mTBI subject.
